# Regional importation and asymmetric within-country spread of SARS-CoV-2 variants of concern in the Netherlands

**DOI:** 10.1101/2022.03.21.22272611

**Authors:** Alvin X. Han, Eva Kozanli, Jelle Koopsen, Harry Vennema, RIVM COVID-19 molecular epidemiology group, Karim Hajji, Annelies Kroneman, Ivo van Walle, Don Klinkenberg, Jacco Wallinga, Colin A. Russell, Dirk Eggink, Chantal B.E.M. Reusken

## Abstract

Variants of concern (VOCs) of SARS-CoV-2 have caused resurging waves of infections worldwide. In the Netherlands, Alpha, Beta, Gamma and Delta variants circulated widely between September 2020 and August 2021. To understand how various control measures had impacted the spread of these VOCs, we analyzed 39,844 SARS-CoV-2 genomes collected under the Dutch national surveillance program. We found that all four VOCs were introduced before targeted flight restrictions were imposed on countries where the VOCs first emerged. Importantly, foreign introductions, predominantly from other European countries, continued during these restrictions. Our findings show that flight restrictions had limited effectiveness in deterring VOC introductions due to the strength of regional land travel importation risks. We also found that the Alpha and Delta variants largely circulated more populous regions with international connections after their respective introduction before asymmetric bidirectional transmissions occurred with the rest of the country and the variant dominated infections in the Netherlands. As countries consider scaling down SARS-CoV-2 surveillance efforts in the post-crisis phase of the pandemic, our results highlight that robust surveillance in regions of early spread is important for providing timely information for variant detection and outbreak control.

## Introduction

Coronavirus-19 disease (COVID-19) has resulted in excess morbidity and mortality across the world. In response, governments have implemented travel restrictions and nonpharmaceutical interventions in order to limit introductions and reduce transmission of severe acute respiratory syndrome coronavirus 2 (SARS-CoV-2)^1–3^. However, high levels of global infections have led to the evolution and emergence of variants of concern (VOCs) that are more transmissible, some of which encode putative mutations that evade immunity acquired from previous infection or vaccination^4^. These VOCs have led to resurging SARS-CoV-2 outbreaks, hampering efforts to contain and control the pandemic worldwide. Of note, four such VOCs arose into global prominence in late 2020, including Alpha (Nextclade 20I; PANGO lineage B.1.1.7), Beta (20H; B.1.351), Gamma (20J; P.1) and Delta (21J; B.1.617.2), causing substantial levels of transmission worldwide, with Alpha and Delta being the most common variants globally in 2021^5^.

The Alpha variant was first reported in the United Kingdom (U.K.) during the fall of 2020 and found to be 43-90% more transmissible^6,7^ with greater mortality risks^8,9^ than previously existing variants. Of the 17 amino acids mutations found in Alpha, N501Y in the receptor-binding domain (RBD) of the spike protein was predicted to increase binding to the human angiotensin-converting enzyme 2 receptors^10^. This is also a common mutation found in Beta^11^ and Gamma^12^. On the other hand, the Delta variant, first identified in India in October 2020^13^, encodes P681R mutation in the furin cleavage site in spike protein and R203M mutation in the nucleocapsid protein that improves infectivity^14^. Delta has also been linked to increased disease severity, as well as greater and longer viral shedding^15^. In the U.K., where the variant was first detected in April 2021, epidemiological modelling estimated the VOC to be 40-80% more transmissible than Alpha.

The four aforementioned VOCs also emerged in the Netherlands, with the Alpha and Delta variants subsequently dominating infections in the country in 2021. In a bid to deter introductions and slow down the spread of VOCs, the Dutch government implemented targeted flight restrictions on countries where these variants had first emerged. Various non-pharmaceutical interventions were also implemented as the country experienced multiple waves of infections between 2020 and 2021. Since the end of 2020, the Dutch National Institute for Public Health and Environment scaled up its sequencing efforts under a random national surveillance program. This detailed surveillance program allows the monitoring of the introduction and spread of novel variants or specific mutations. Not only would such data help characterize source-sink dynamics in order to assess importation risks of novel variants and elucidate within-country transmission dynamics, genomic epidemiology can also help shed light on the impact of virus transmission control and relaxation strategies. Here, 39,844 high-quality SARS-CoV-2 whole genomes were randomly collected across the country and sequenced between 22 September 2020 and 31 August 2021 (48 calendar weeks) to characterize the importation risks and spread of novel SARS-CoV-2 variants in the Netherlands.

## Results

### SARS-CoV-2 infections and genotypes circulating in the Netherlands from September 2020 to August 2021

There were 1,792,759 laboratory-confirmed SARS-CoV-2 cases in the Netherlands during the study period between 22 September 2020 and 31 August 2021 (week 39/2020 to week 34/2021; Figure 1A). Similar to the first wave of the pandemic in the Netherlands in Spring 2020, most reported cases were attributed to the more densely populated regions of the country including North and South Holland, as well as North Brabant (Figure 1B) where the first local clusters of SARS-CoV-2 were also detected in March 2020^16^. 39,844 SARS-CoV-2 positive nasopharyngeal samples were randomly selected from 25 Municipal and Regional Health Service (GGD) regions across the Netherlands during this study period and sequenced to obtain whole virus genomes as part of the national SARS-CoV-2 genomic surveillance program.

**Figure 1:**
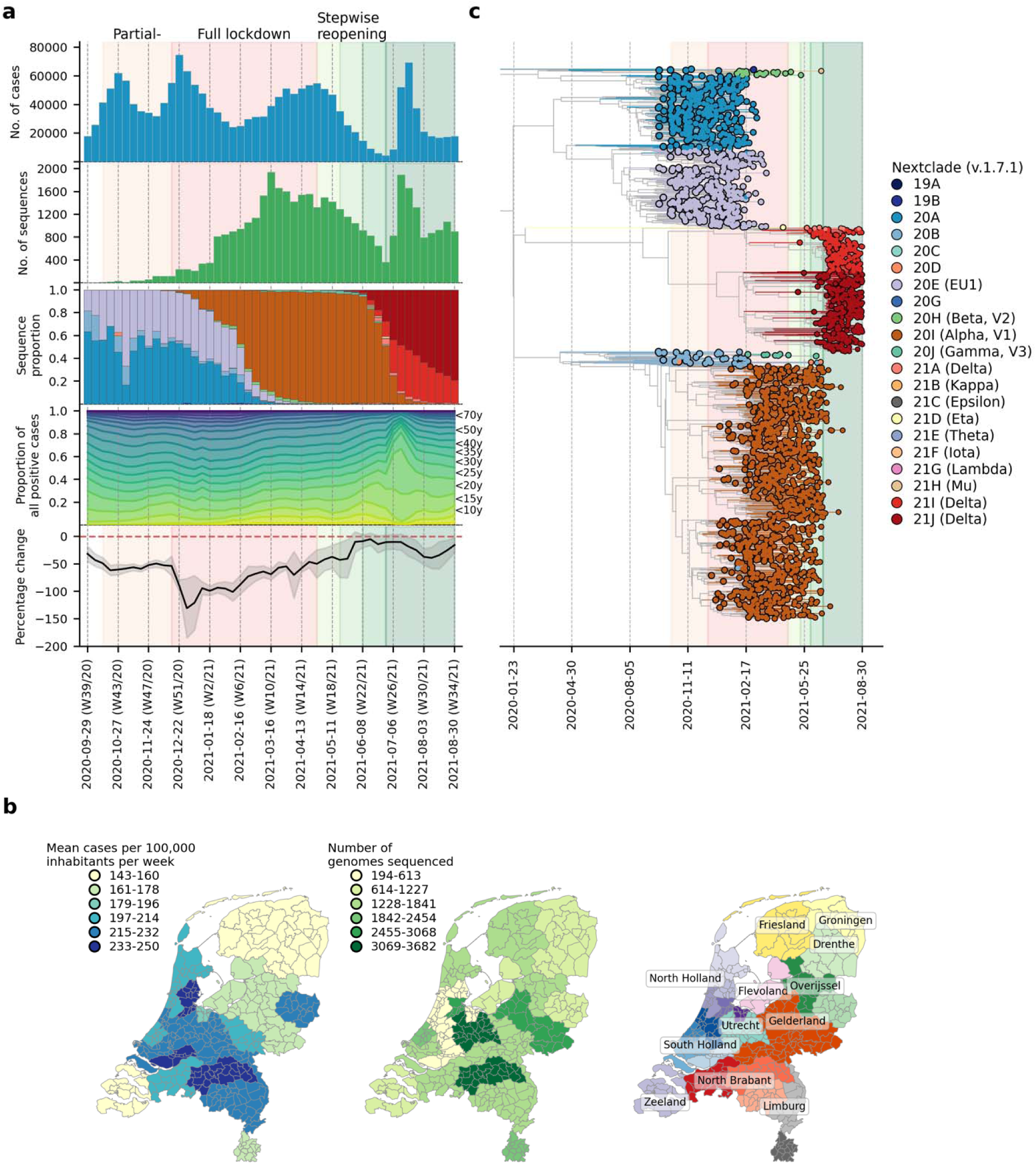
SARS-CoV-2 infections and genotypes circulating in the Netherlands from September 2020 to August 2021. (**A**) Weekly number of laboratory-confirmed SARS-CoV-2 cases (1st panel from the top) and sequenced genomes (2nd panel). Genotype proportions of sequences (3rd panel) are shown as stacked bars colored by NextClade designations as in (C). Breakdown of positive cases by age group from data provided by the Dutch National Institute for Public Health and Environment (4th panel). Aggregated weekly average percentage change in mobility to the baseline in the Netherlands from Google’s COVID-19 community mobility reports. Baseline mobility is the median value from a 5-week period between 3 January 2020 and 6 February 2020, prior to the COVID-19 pandemic in Europe (5th panel). (**B**) Mean number of laboratory-confirmed cases per 100,000 inhabitants (data from the Dutch National Institute for Public Health and Environment) and total number of sequenced genomes in different Municipal and Regional Health Service (GGD) regions over the entire study period. (**C**) Maximum clade credibility tree of sequences based on a downsampled set of 2,246 sequences that is representative of the weekly relative proportions of reported case numbers across different GGD regions. Tips are colored by NextClade genotype designations.

Using NextClade lineage assignment^17^, the viruses sampled at the start of the study period were largely genotyped as clade 20A and its daughter lineages, 20B and 20E (EU1) (Figure 1B-C). 20A was the lineage that seeded the pandemic in Europe in March 2020. On the other hand, 20E (EU1) was first detected in Spain on June 2020 and spread widely across Europe due to the resumption of regional travel over summer 2020^18^. Owing to rising case numbers, non-pharmaceutical interventions closing restaurants and nightlife establishments were implemented on 14 October 2020. Cases dipped momentarily while both 20A and 20E (EU1) remained co-circulating into December 2020.

The first Alpha sample was collected on 5 December 2020 in the national surveillance program prior to the full lockdown that closed all public venues, workplaces and schools on 15 December 2020. A curfew was also imposed later on 23 January 2021. A sharp drop in cases was observed after the implementation of the full lockdown. Alpha then displaced 20A and 20E (EU1) over time to become the dominant circulating virus lineage by 16 February 2021 (week 6) for the rest of the lockdown period. Other VOCs such as Beta (N=422 sequences; first sequence was collected on 22 December 2020) and Gamma (N=350 sequences; first sequence was collected on 27 January 2021) were also detected by random surveillance in The Netherlands around the turn of the year but did not circulate to the levels of Alpha.

Alpha caused a rebound in cases around mid-March 2021, after which case numbers stabilized and eventually began to decline at the end of April 2021. The Dutch government began taking steps to relax restrictions around the same time, starting with the end of curfew and resumption of higher education during the week of 27 April 2021. The first Delta sample was collected in the previous week on 15 April 2021 and continued to accumulate in frequencies. By week 25 (29 June 2021), the Delta variant accounted for 24% of all weekly genomes sequenced. Most restrictions were lifted in the same week including the reopening of nightlife establishments on 26 June 2021. SARS-CoV-2 prevalence was at its lowest then with only 8,690 reported positive cases that week. Within only one week after reopening, however, weekly cases soared above 50,000 on weeks 26 and 27 (6-20 July 2021). With most infections attributed to Delta, the novel VOC replaced the Alpha variant as the dominant lineage within the next three weeks as over 90% of randomly surveilled genomes were typed as Delta variants by mid-July.

Stratifying the number of weekly reported positive cases by patient age group, the relative proportions in case positive rates remained fairly consistent throughout the study period except for weeks 26 and 27 where the rapid increase in cases was largely attributed to individuals aged between 15-30 years (Figure 1A). One of the reasons behind widespread transmission among young adults then was super-spreading linked to nightlife venues^19^. In response, the government shut nightlife establishments down again on 10 July 2021 (week 27). Case numbers fell promptly after but remained at over 30,000 new cases per week for the rest of the study period. The Delta variant had in principle completely displaced Alpha by then with over 99% of randomly surveilled genomes sampled from August 2021 onwards.

### Overseas introduction of variants of concern

To understand where and when VOCs were introduced into the country, we subsampled a representative set of Dutch and overseas sequences collected over the same time period. We then reconstructed time-scaled, maximum likelihood (ML) phylogenies and used these fixed trees to perform discrete trait analyses using a Bayesian approach to infer likely overseas introductions at the continental level. This was done by identifying subtrees subtending Dutch sequences with ancestral states that were attributed to an overseas origin (Figure 2). For all four VOCs, the variant viruses were already introduced into the Netherlands prior to the targeted flight restrictions that were imposed on countries where these variants first emerged.

**Figure 2:**
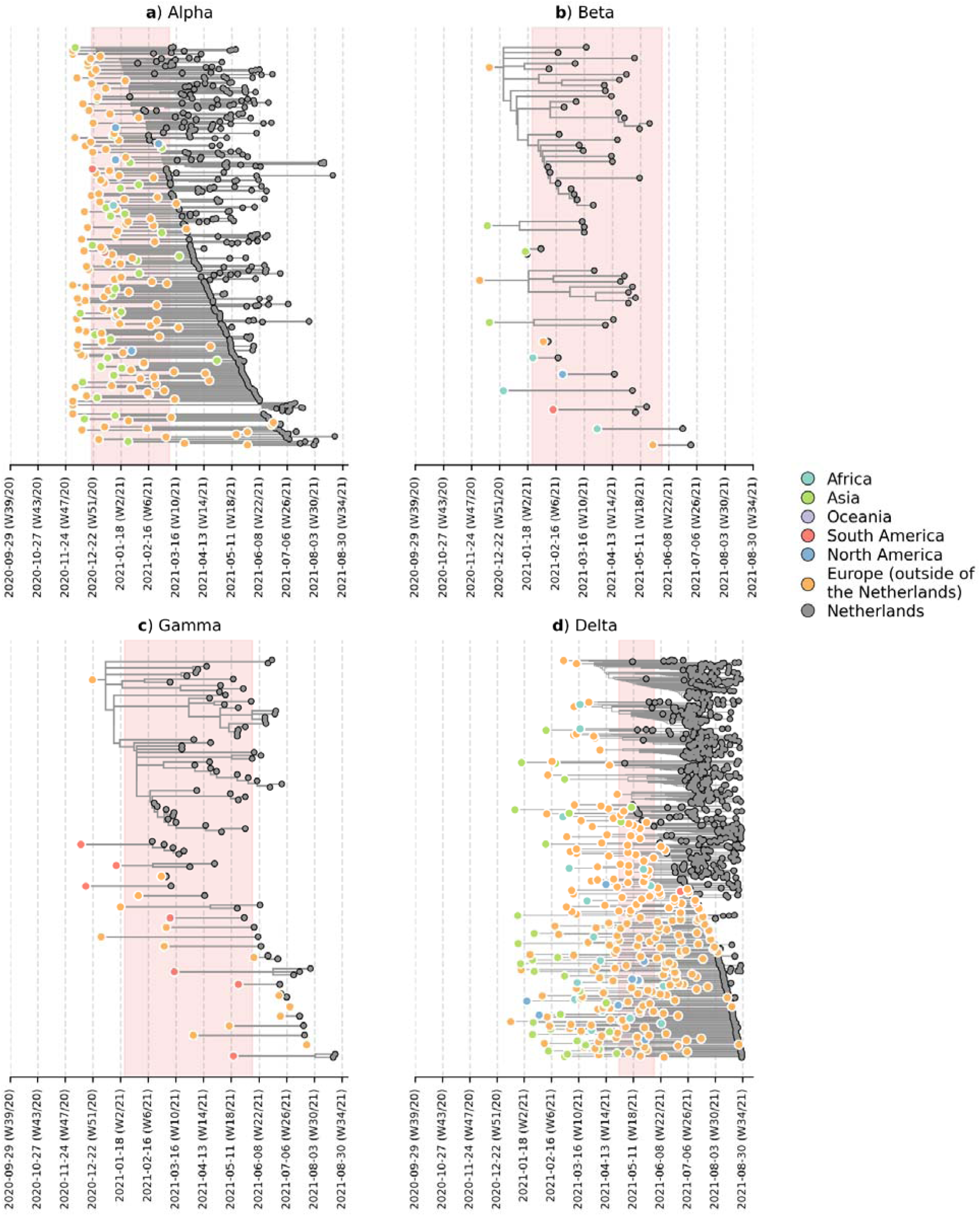
Likely overseas introduction of VOC lineages into the Netherlands at the continental level. For each VOC lineage, a time-scaled maximum likelihood phylogeny using the Dutch and their nearest overseas neighboring sequences was inferred. Discrete trait analyses were performed to infer the likely continental region of ancestral states. Subtrees or singletons with ancestral nodes attributed to an overseas origin but subtend only Dutch sequences are drawn. Shaded plot area denotes the timespan when a targeted flight restriction was imposed on the country where the VOC lineage first emerged (i.e. (**A**) Alpha, United Kingdom.; (**B**) Beta, South Africa; (**C**) Gamma, Brazil; (**D**) Delta, India)

Importantly, besides countries where travel restrictions were in place, we estimated multiple likely introduction events from other foreign countries into the Netherlands for all four VOCs (Alpha, n=100; Beta, n=7; Gamma, n=12; Delta, n=213). Given disparities in global sequencing efforts^20^, the random surveillance strategy used in local sample collection, and low genetic diversity among SARS-CoV-2 genomes used to reconstruct ancestral states, we are unable to fully and reliably quantify the number of introductions attributed to different geographical regions. However, many of the estimated regions for these ancestral states were in Europe (Alpha, 71% of all estimated overseas introduction events; Beta, 29%; Gamma, 71%; Delta, 79%; Figure 2). Furthermore, these European introductions continue to occur during the targeted travel ban period. Inspecting the nearest phylogenetic ancestral taxon to the aforementioned subtrees, we found that many of these nearest overseas neighboring tips were detected in Belgium, Germany, France and Denmark where land borders between the Netherlands remained open as well as other countries (e.g. Spain, U.K., Poland, U.S.) where no targeted travel restrictions were set in place (Figure 2 – figure supplement 1). There was also no isolated period in time in which these VOCs were introduced into the Netherlands - introductions likely occurred repeatedly during the period when these variants were also proliferating within the country.

### Within-country transmission dynamics of the Alpha and Delta variants

To further elucidate the transmission dynamics of the Alpha and Delta variants within the Netherlands, we performed continuous phylogeographic analyses using separate downsampled sets of Alpha and Delta sequence data (Figure 3). For the first four weeks since the initial detection of both variants within the country, introductions and phylogenetic branch movements were mostly concentrated in the more populous regions of the country, including North and South Holland, Utrecht and North-Brabant (Figure 3A, 3C and 4A), forming a core of early dominant locations. During this period, dispersal events to regions outside of these GGD regions occurred as well but are relatively less frequent. However, as local infections were seeded in these areas, bidirectional exchanges in phylogenetic branches between different regions emerged throughout the country. These bidirectional exchanges continued to increase as prevalence of the variant grew over time, even amidst a strict lockdown in the case of Alpha (Figure 3A-B and 4A). In particular for the Delta variant, we observed a rapid spike in inter-regional spread upon the week of nightlife reopening (22-28 June 2021), with >400% estimated increase in total phylogenetic branch movements by 6 July 2021 (week 26) (Figure 3C-D and 4A). This significant rise in inter-regional exportation events likely contributed to the soaring case numbers observed between weeks 25 and 27 (22 June – 13 July 2021).

**Figure 3:**
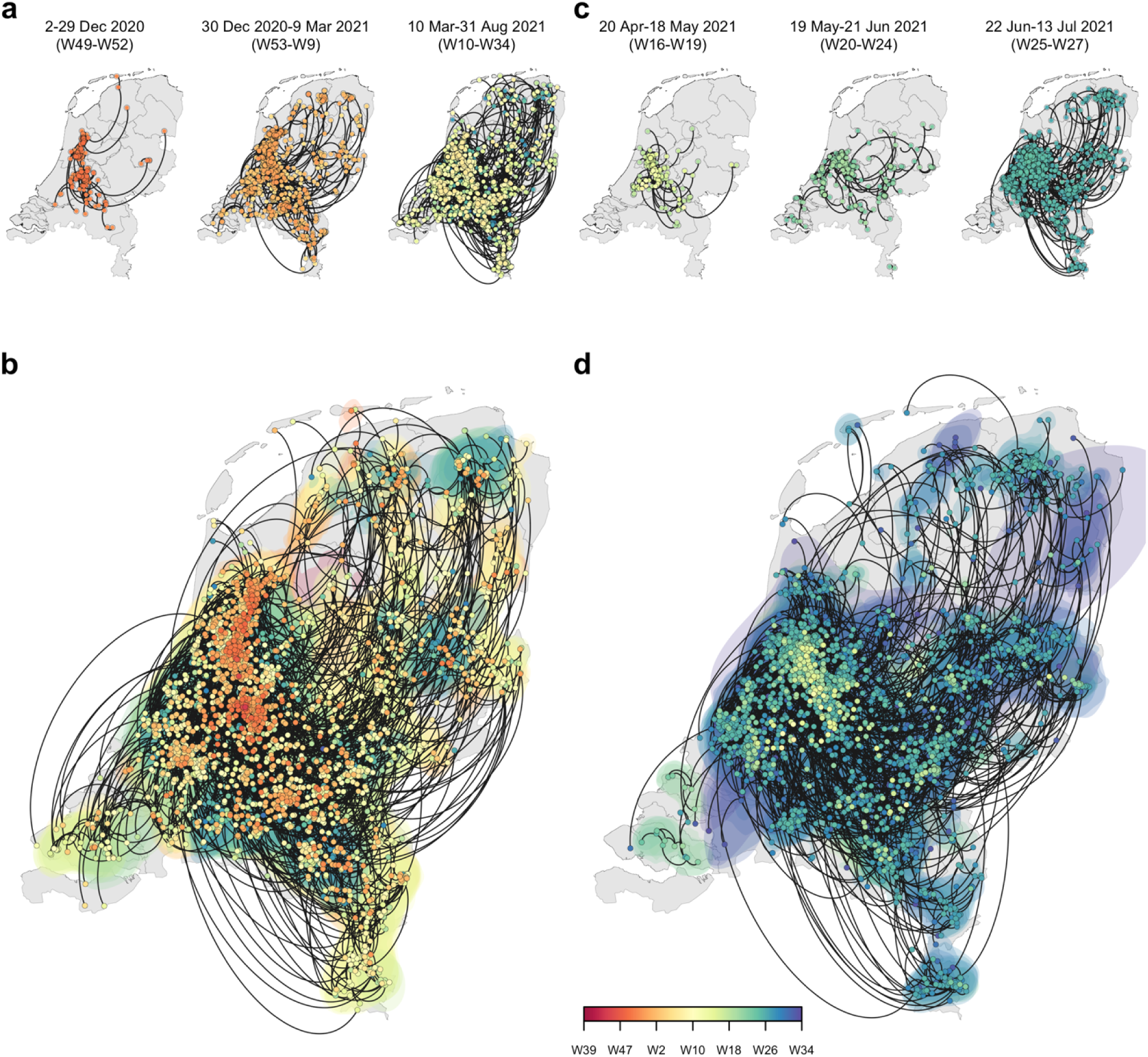
Spatiotemporal spread of the Alpha and Delta variants of concern in the Netherlands. Reconstruction of continuous phylogeography with nodes colored by time and the dispersal directionality of phylogeny branches with counterclockwise edges. (**A, B**) Alpha variant; (**C, D**) Delta variant. Top panels (A) and (C) show the reconstruction of specific labelled periods for each variant. Bottom panels (B and (D) show the reconstruction for the entire study period.

We also fitted the observed weekly variant proportions of sampled viruses to a logistic growth model to quantify the growth rates of both VOCs in the Netherlands. For the Delta variant, we found a weekly growth rate of 103% (95% CI = 97-110%), which was about two times higher than Alpha, with a weekly growth rate of 45% (95% CI = 40-51%) (Figure 4B). Given previous reports of superspreading events among young adults after nightlife reopening^19^ and the relatively higher case numbers attributed to these age groups (15-30 years) in weeks 26-27 (Figure 1A), we repeated the logistic regression for age-stratified weekly proportion data for both variants to assess if there were differences in growth rates between age groups as well. For Alpha, estimated growth rates were similar across ages (weekly growth rates 35-46%), but for Delta we observed a slightly increased weekly growth rate in the 30-39 age group (122% versus 92-111% for other age groups). Nonetheless, for both Alpha and Delta variants, we estimated that dominance was reached for different age groups at similar times (by week 5-7 (between 3 and 23 February 2021) for Alpha, by week 25-26 (between 23 June and 6 July 2021) for Delta, Figure 4 – figure supplement 1, Figure 4 – table supplement 1). Both Alpha and Delta variants replaced previous variants rapidly and with similar rates across most age groups. In other words, while there was a greater increase in cases among young adults after nightlife reopening, the Delta variant did not displace Alpha more rapidly in these age groups only (Supplementary Table S1).

**Figure 4:**
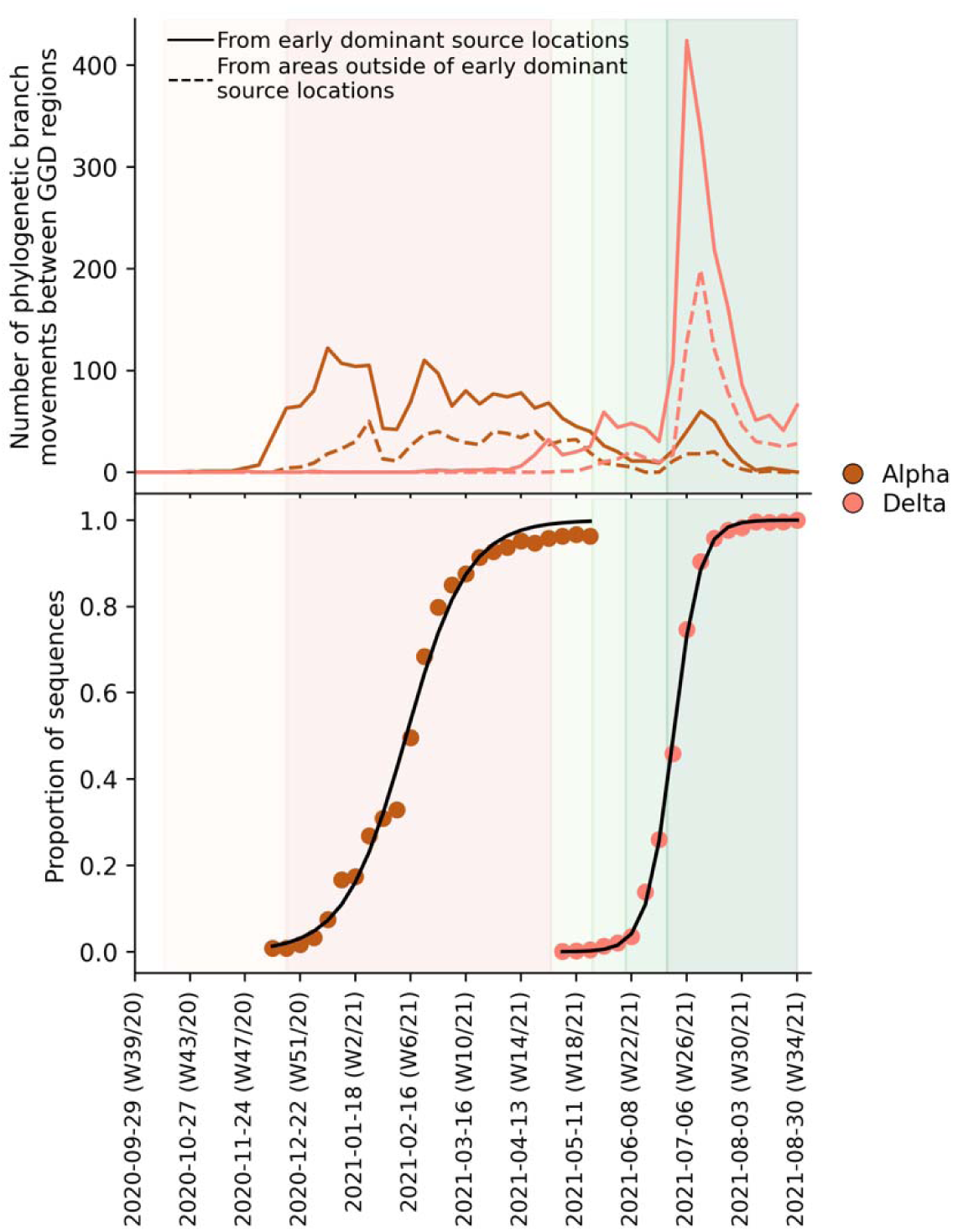
Estimated number of phylogenetic branch movements and growth rate estimations of Alpha and Delta variants in the Netherlands. (**A**) Estimated number of phylogenetic branch movements between GGD regions over time as shown in Figure 3B and D. Solid line shows the number of branch movements from early dominant source regions including North and South Holland, Utrecht and Brabant. Dashed line shows number of branch movements from areas outside of these early dominant source locations. (**B**) Logistic growth regression of Alpha and Delta sequence proportions.

## Discussion

Even if international travel restrictions are in place, the Netherlands is still highly vulnerable to importation risks of novel SARS-CoV-2 variants from its regional neighbors due to border policies within the European Union. As such, this regional vulnerability is not unique to the Netherlands and has been reported in other European countries as well^21–23^. Importantly, regional introductions of novel lineages often drive new waves of infections in Europe^23^. Prior to September 2020, the dominant variant lineages (i.e. 20A and 20E (EU1)) that circulated the Netherlands were already seeded by imports from its European neighbours^16,18,24^. In fact, the initial introduction of SARS-CoV-2 in the Netherlands in February 2020 were attributed to travelers who visited Northern Italy where the earliest sustained European SARS-CoV-2 transmission network was seeded^16,25^. Here, we showed that all four VOC lineages detected in the country up to August 2021 also originated mainly from its European neighbors. Importantly, regional importation risks persisted throughout the period these variants circulated the country and overlapped with periods where targeted flight restrictions were imposed on countries where these VOCs first emerged. The recent emergence of the Omicron variant in southern Africa in November 2021^26^ again led to reactionary targeted flight restrictions by several countries in the Global North, including the Netherlands which was still amidst a surging Delta infection wave. However, the Omicron variant was already detected in samples collected one week before the imposed flight ban and did not prevent it to rapidly become the dominant variant circulating in the Netherlands by the end of 2021 (https://www.rivm.nl/coronavirus-covid-19/virus/varianten/omikronvariant). Previous studies showed that travel restrictions are only useful if restrictions barred arrivals from most countries provided that local incidence is low in the first place^27^.

We also found that early introductions of VOCs, specifically the Alpha and Delta variants, are more likely found in populous regions of the Netherlands, including Utrecht, North and South Holland where larger cities are locatthed that are also international and regional travel hubs. These areas constitute a core cluster of dominant source locations that also exported infections to the rest of the country during first few weeks after the VOC’s introduction into the Netherlands. As the number of infections in areas outside of these dominant source locations increase over time, bidirectional exchanges would also become more frequent. This type of asymmetric spatial spread dynamics had been previously shown in the U.K. as well and was found to enhance the intrinsic transmissibility of Alpha^28^. Additionally, enhanced mobility has also been previously linked to the resurgence of outbreaks across Europe^18,23^. Recent work also showed that increased mobility and population mixing drove the rapid dissemination of Delta in the U.K.^29^. In the case of the Netherlands, the declining prevalence of SARS-CoV-2 after the hard lockdown since mid-December 2020 was mirrored by an increase in average nationwide mobility that eventually came close to pre-pandemic levels by June 2021 before nightlife reopening (mean percentage change relative to pre-pandemic baseline = -5.0% (s.d. = 11.0%); Figure 1A). While our analyses do not provide a causal relationship between the relaxation of non-pharmaceutical interventions and frequency of export events, the asymmetric exportation frequencies from dominant source locations, increased human mobility in the country as well as the intrinsic higher transmissibility of Delta relative to Alpha likely all contributed to the widespread spike in cases in weeks 25-27 across the country.

Novel and fitter variants of SARS-CoV-2 will likely continue to emerge in the future. Our results, along with others, show that unless well-coordinated actions are taken across Europe to mitigate importation risks^30^, targeted travel restrictions implemented by individual European countries will neither prevent nor slow down the introduction of novel variants. Our work also shows that early within-country spread of VOCs may be taken into future consideration in future genomic surveillance strategies, especially as countries are gradually considering scaling down SARS-CoV-2 surveillance efforts. Both the Alpha and Delta variants were first detected in the early dominant source locations, usually those that are more populous with greater international connections, and circulated mostly within these areas during the initial period after introduction. As such, a robust level of surveillance efforts should still be maintained in these dominant source locations to provide timely actionable information on novel variant detection as well as infection control.

## Methods

### Whole genome sequencing

39,844 nasopharyngeal samples were randomly collected across all 25 GGD health services across the Netherlands and were sequenced for whole SARS-CoV-2 genomes. Amplicon-based SARS-CoV-2 sequencing for was performed using the Nanopore protocol “PCR tiling of COVID-19 virus (Version: PTC_9096_v109_revE_06FEB2020)” which is based on the ARTIC v3 amplicon sequencing protocol^31,32^. Several modifications were made to the protocol as primer concentrations were increased from 0.125 to 1 pmol for the following amplicon primer pairs: in pool A amplicons 5, 9, 17, 23, 55, 67, 71, 91, 97 and in pool B amplicons 24, 26, 54, 64, 66, 70, 74, 86, 92, 98. Both libraries were generated using native barcode kits from Nanopore SQK-LSK109 (EXP-NBD104, EXP-NBD114 and EXP-NBD196) and sequencing was performed on a R9.4.1 flow cell multiplexing 2 up to 96 samples per sequence run.

### Epidemiological data

All epidemiological data including the breakdown of positive cases by age group and weekly number of laboratory-confirmed cases in each Municipal and Regional Health Service region are provided by the Dutch National Institute for Public Health and Environment (https://www.rivm.nl/en/node/163991).

### Phylogenetic analyses

We downsampled the full dataset of SARS-CoV-2 Dutch genomes to a representative set of 2,246 sequences. This was done by randomly subsampling the number of sequences in each GGD region each week to the corresponding relative number of reported cases in the same week for that GGD region. All sequences were aligned to hCoV-19/Wuhan/WIV04/2019 (WIV-04; EPI_ISL_402124) using MAFFT v7.427^33^. Likely problematic sites (https://github.com/W-L/ProblematicSites_SARS-CoV2) along with untranslated regions in the 5’ and 3’ ends were masked. For each downsampled set of data, a maximum-likelihood (ML) phylogenetic tree was reconstructed using IQ-TREE^34^ under the Hasegawa–Kishono–Yano nucleotide substitution model with a gamma-distributed rate variation among sites (HKY+G). We regressed the root-to-tip genetic distances against sampling dates using treetime v0.8.1^35^ to assess the level of temporal signal, ensuring that none of the representative sequences were deemed molecular clock outliers. We then reconstructed a time-resolved phylogeny using BEAST v1.10.4^36^ under the HKY+G nucleotide substitution model and a Skygrid coalescent model^37^ (each grid point denoting one week) with a strict molecular clock. Due to the lack of a strong temporal signal, we used an informative clock prior (Gamma distribution with *µ* = 8 × 10^−4^ substitutions/site/year and *σ* = 5 × 10^−4^) similar to those used in other recent phylogenetic analyses of SARS-CoV-2 that reflect the latest estimates of its substitution rate^38^. The respective molecular clock rooted ML tree inferred was used as the starting tree. We performed 300 million MCMC generations that were sampled every 50,000 steps. The first 100 million steps were discarded as burn-in. Assessment of convergence (effective sample size > 200) was performed using Tracer v1.71^39^.

The aforementioned procedure, with the exception of subsampling equitably over all GGD regions in each week, was also used to obtain downsampled sets of Alpha (n=1,389) and Delta (n=1,342) variant sequences collected in the Netherlands. To understand within-country source-sink dynamics during early introductions and proliferation patterns during later periods, we used BEAST v.1.10.4 to perform continuous phylogeographical analyses on these sequence data, using a relaxed random walk diffusion model and a Cauchy distribution model among branch heterogeneity in diffusion velocity^40^. We inferred geographical coordinate input using the first four digits of postcodes (i.e. neighborhood level) associated with the sampled sequences. For sequences with identical postcodes, we randomly selected geographical coordinates corresponding to the postcode area using shapefiles provided by https://www.gadm.org. Similarly, we performed 300 million MCMC generations for each variant analysis, sampling every 50,000 steps. Visualization was performed using customized scripts from the SERAPHIM package^41^.

We also performed ancestral reconstruction analyses for each VOC lineage to identify likely overseas introduction into the Netherlands at the continental level, differentiating the Netherlands from the rest of Europe. As proportions of cases for each VOC lineage are unknown for most countries, we subsampled global sequences downloaded from GISAID (https://www.gisaid.org; dataset up to 6 October 2021) by the proportion of COVID-19 cases reported per week for each country using data from the Johns Hopkins University, Center for Systems Science and Engineering (CSSE) (http://github.com/CSSEGISandData/COVID-19). We sampled 100 global sequences each week, ensuring that at least one representative sequence was included for each country with reported cases that week. We also subsampled Dutch sequences based on the weekly number of cases in different GGD regions as described above and strived to maintain a 2:1 sampling ratio between global and Dutch sequences. The subsampling procedure yielded 6,365 (2,369), 1,531 (90), 1,274 (102) and 6,929 (1,035) Alpha, Beta, Gamma and Delta global (Dutch) sequences respectively. Using these sequences, we then reconstructed approximate ML phylogenies using FastTree v2.1.11^42^. All phylogenetically neighboring overseas sequences placed within two nodes away from any Dutch sequence were retained. This further reduced the number of sequences to a representative set of 3,671, 496 and 575 and 2,180 sequences for Alpha, Beta, Gamma and Delta variants respectively. Once again, we aligned and masked problematic sites for this downsampled set of Dutch and overseas sequences. Similarly, we reconstructed an ML phylogenetic tree under the HKY+G nucleotide substitution model using IQ-TREE after removing any molecular clock outlying sequences identified by treetime. Here, however, we included the WIV-04 reference genome in the phylogeny reconstruction which was used as an outgroup for tree rooting. We then time-scaled these ML phylogenies using treetime, which were then used as fixed tree topologies in BEAST v.1.10.4 to perform Bayesian discrete phylogeographical analyses at the continental level. Here, we performed 100 million MCMC generations, sampling every 1,000 steps.

All tree visualizations were performed using baltic (https://github.com/evogytis/baltic).

### Aggregated mobility data

We used publicly available mobility data from Google COVID-19 community mobility reports (https://www.google.com/covid19/mobility/) which contain daily anonymized location histories as a measure of people’s movements. Google mobility data consisted of six categories that were measured relative to a baseline value. This baseline is the median mobility value between pre-pandemic weeks of 3 January and 6 February 2020. Categories include residence, parks, retail and recreation, groceries and pharmacies, working place and transit. Data for different regions of the Netherlands were available. We calculated aggregated nationwide mean mobility by averaging values across all regions for all categories except for residence and parks where the former has a reversed effect on relative mobility while the latter is affected by climate.

### Relative growth rate estimation of the Alpha and Delta variants

We used the *nlsLM* function of the *minpack*.*lm* package^43^ in *R* to fit a logistic growth model to aggregated, weekly proportions of sequences genotyped to Alpha and Delta.

## Data availability

All codes for our analyses are available at https://github.com/AMC-LAEB/nl_sars-cov-2_genomic_epi_2022.

## Acknowledgements

We thank the administrators of the GISAID database for supporting rapid and transparent sharing of genomic data during the COVID-19 pandemic and all our colleagues sharing data on GISAID. A full list acknowledging the authors submitting genome sequence data used in this study can be found in Supplementary File 1. AXH and CAR were supported by ERC NaviFlu (no. 818353). C.A.R. was also supported by NIH R01 (5R01AI132362-04) and an NWO Vici Award (09150182010027).

## Supplementary material

**Figure 2 – figure supplement 1.**
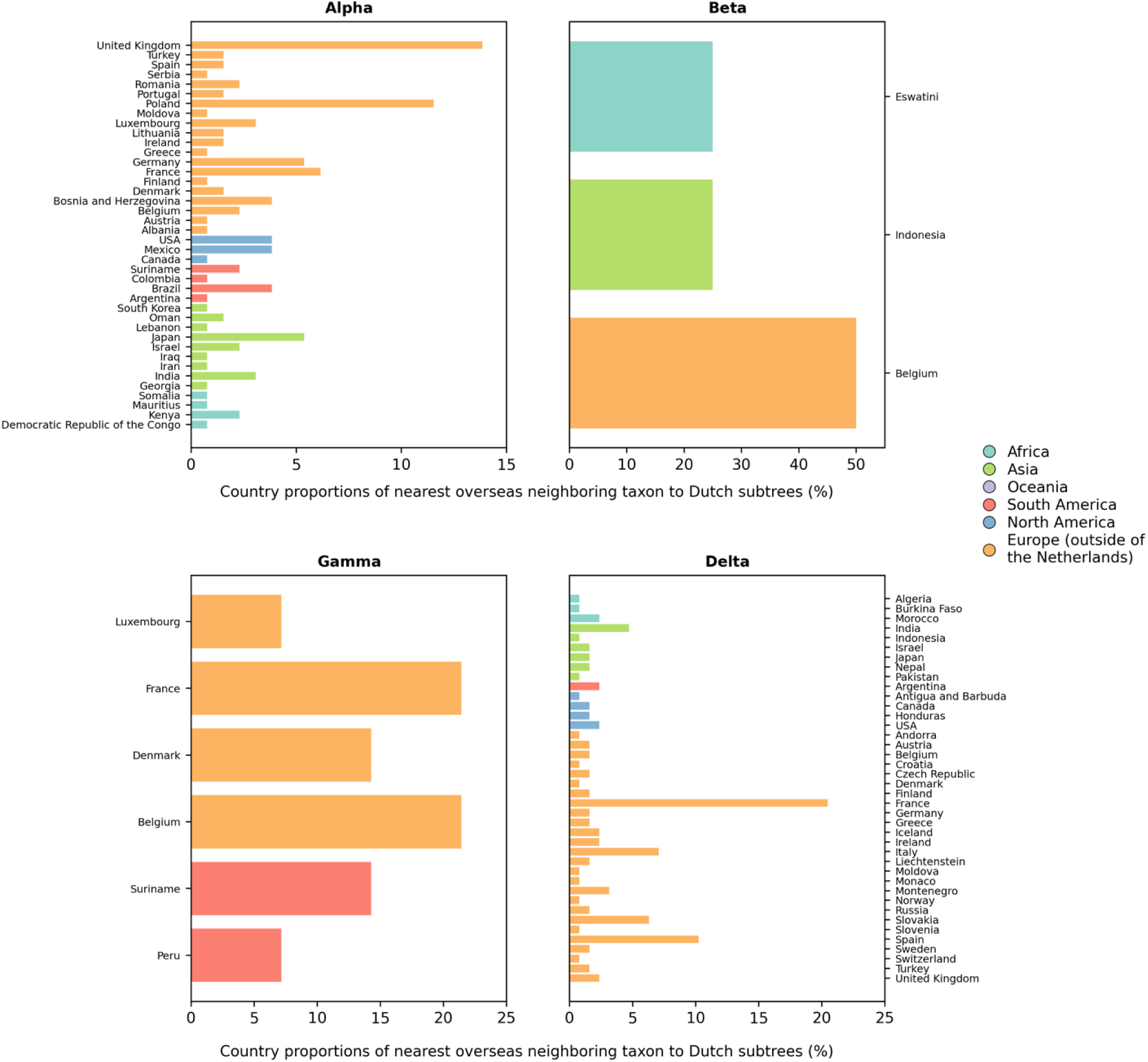
Distribution of countries of the nearest overseas neighboring taxon to Dutch subtrees.

**Figure 4 – figure supplement 1.**
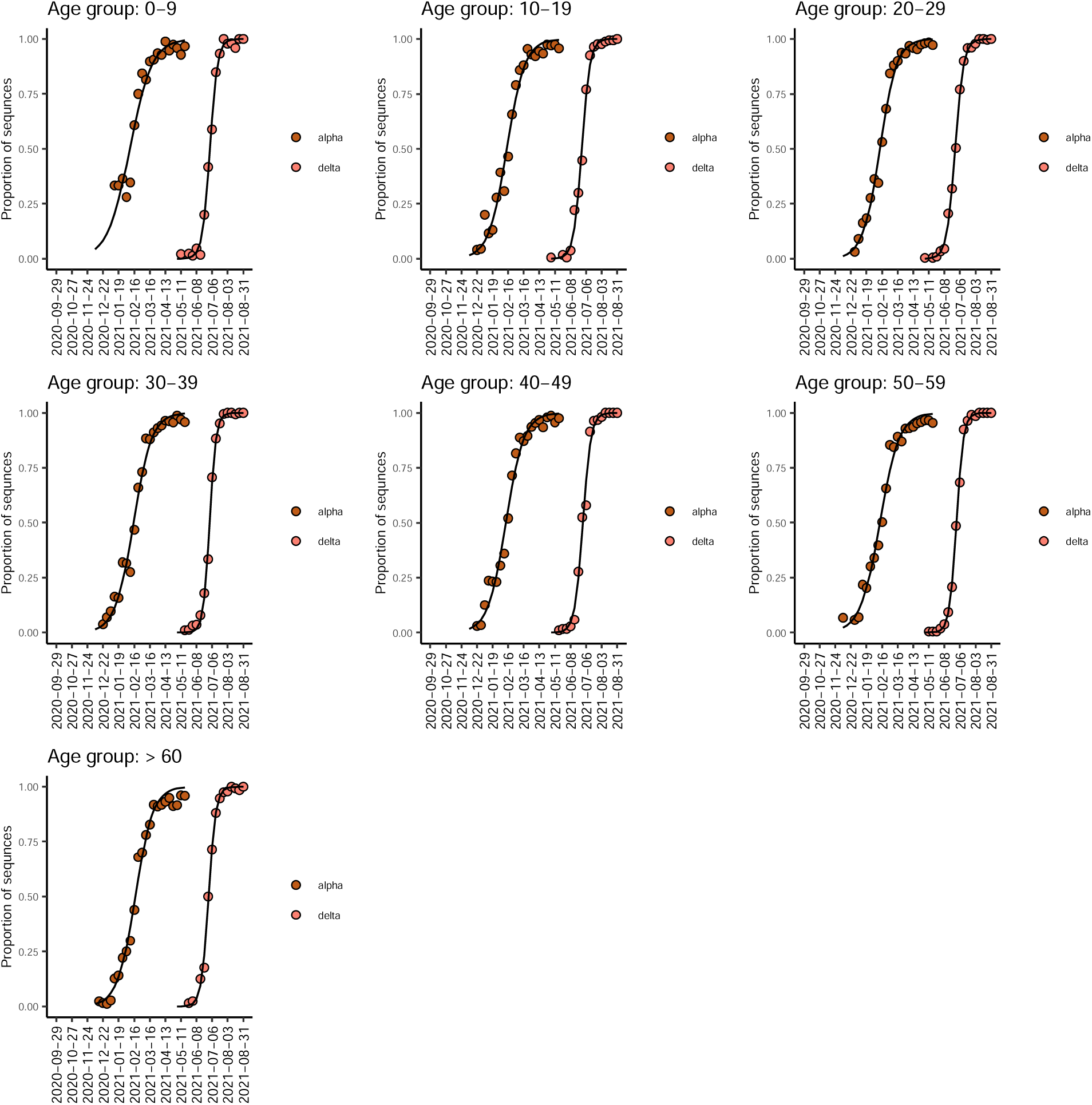
Logistic growth regression of Dutch Alpha and Delta sequence proportions stratified by patients’ age.

**Figure 4 – table supplement 1.**
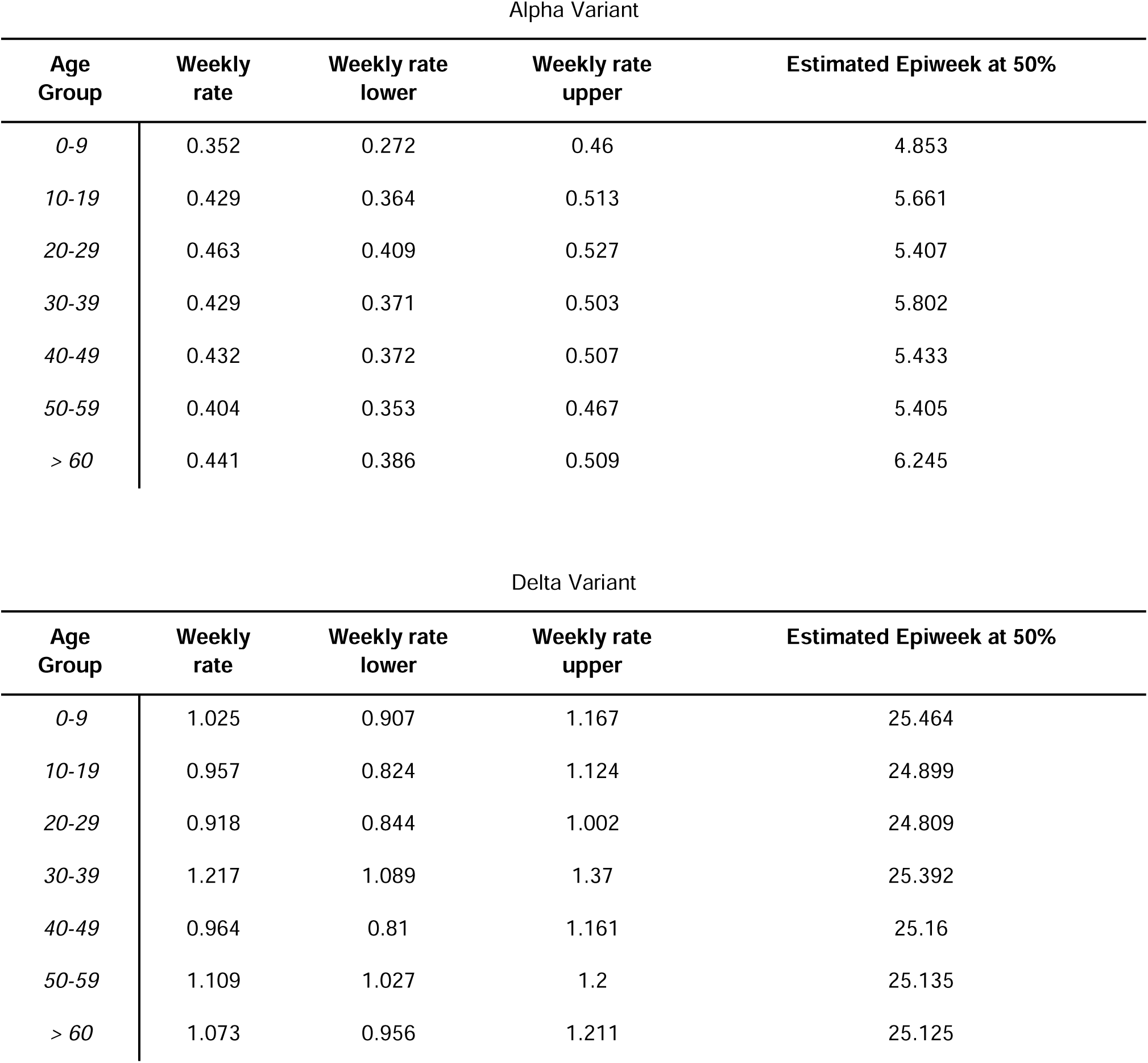
Weekly growth rates per age group of Alpha and Delta variant. * Defined as timepoint (week of the year) where the proportion of the variant reached 50%.

| Alpha Variant |  |  |  |  |
| --- | --- | --- | --- | --- |
| Age Group | Weekly rate | Weekly rate lower | Weekly rate upper | Estimated Epiweek at 50% |
| 0-9 | 0.352 | 0.272 | 0.46 | 4.853 |
| 10-19 | 0.429 | 0.364 | 0.513 | 5.661 |
| 20-29 | 0.463 | 0.409 | 0.527 | 5.407 |
| 30-39 | 0.429 | 0.371 | 0.503 | 5.802 |
| 40-49 | 0.432 | 0.372 | 0.507 | 5.433 |
| 50-59 | 0.404 | 0.353 | 0.467 | 5.405 |
| > 60 | 0.441 | 0.386 | 0.509 | 6.245 |

| Delta Variant |  |  |  |  |
| --- | --- | --- | --- | --- |
| Age Group | Weekly rate | Weekly rate lower | Weekly rate upper | Estimated Epiweek at 50% |
| 0-9 | 1.025 | 0.907 | 1.167 | 25.464 |
| 10-19 | 0.957 | 0.824 | 1.124 | 24.899 |
| 20-29 | 0.918 | 0.844 | 1.002 | 24.809 |
| 30-39 | 1.217 | 1.089 | 1.37 | 25.392 |
| 40-49 | 0.964 | 0.81 | 1.161 | 25.16 |
| 50-59 | 1.109 | 1.027 | 1.2 | 25.135 |
| > 60 | 1.073 | 0.956 | 1.211 | 25.125 |

